# Predictors of Developmental Defects of Enamel in the Primary Maxillary Central Incisors using Bayesian Model Selection

**DOI:** 10.1101/2022.04.07.22273577

**Authors:** Susan G. Reed, Sijian Fan, Carol L. Wagner, Andrew B. Lawson

**Affiliations:** Department of Pediatrics, Darby Children’s Research Institute, Medical University of South Carolina, Charleston, SC; Department of Statistics, University of South Carolina, Columbia, SC; Department of Public Health Sciences, Medical University of South Carolina, Charleston, SC

**Keywords:** Developmental Defects of Enamel, DDE, Enamel, Hyperplasia, Opacities, Post-eruptive Breakdown, Bayesian Analysis, Gibbs Variable Selection, Linear and Fractional Polynomial Models, Gestational Age, 25 Hydroxyvitamin D, Parathyroid Hormone

## Abstract

Localized non-inheritable developmental defects of tooth enamel (DDE) are classified as enamel hypoplasia (EH), opacity (OP) and post-eruptive breakdown (PEB) using the Enamel Defects Index. To better understand the etiology of DDE, and in particular possibly modifiable variables, we assessed the linkages amongst exposome variables during the specific time duration of the development of the DDE. In general, the human primary central maxillary incisor teeth develop between 13-14 weeks *in utero* and 3-4 weeks’ postpartum of a full-term delivery, followed by tooth eruption at about 1 year of age. We utilized existing datasets of mother and child dyad data that encompassed 12 weeks’ gestation through birth and early infancy, and child DDE outcomes from digital images of the erupted primary maxillary central incisor teeth. We applied a Bayesian modeling paradigm to assess the important predictors of EH, OP, and PEB. The results of Gibbs variable selection showed a key set of predictors: mother’s pre-pregnancy body mass index (BMI); maternal serum levels of calcium and phosphorus at gestational week 28; child’s gestational age; and both mother’s and child’s functional vitamin D deficiency (FVDD). In this sample of healthy mothers and children, significant predictors for OP included the child having a gestational period > 36 weeks and FVDD at birth, and for PEB included a mother’s pre-pregnancy BMI < 21.5 and higher serum phosphorus level at week 28.

## 1. Introduction

Three major types of localized non-inheritable developmental defects of tooth enamel (DDE) are classified in the Enamel Defects Index as enamel hyperplasia (EH), opacities (OP) and post-eruptive breakdown (PEB) (1). Prevalence data for these defects in the primary dentition are scarce, and the results generally reflect global convenience samples with ranges of 4-99%, 2-98% and 6-50% for the three defects, respectively (2–6). One of the consequences that these developmental defects can incur is dental treatment. For example, the enamel surface irregularities of EH can provide niches for cariogenic bacteria leading to dental caries and the subsequent need for dental treatment (7, 8). Dental treatment in these young children with DDE is challenging and expensive (9–13); and 50% of the early childhood dental caries are found in the primary maxillary anterior dentition (14). For OP, the treatment considerations generally revolve around esthetics; and for PEB, the treatment considerations can involve function and/or esthetics (14–16). Prevention efforts for DDE are hampered by the lack of knowledge of potentially modifiable etiological variables during the development of the structurally defective enamel.

To address this need for knowledge of the variables and their interplay during the development of EH, OP and PEB, we focused our study on DDE in the human primary central maxillary incisor teeth. The crowns of these two teeth develop between 13-14 weeks’ *in utero* and 3-4 weeks’ postpartum of a full-term delivery (17, 18). To encompass the developmental period for these teeth, we assessed pertinent variables from the mother during pregnancy and at delivery; and the child at delivery through 4-6 weeks of infancy. The outcomes were determined from digital images of the erupted primary maxillary incisor teeth. We conducted a secondary analysis of maternal and child data from a randomized controlled clinical trial (RCT) of vitamin D supplementation for healthy mothers during pregnancy and birth, and child follow-up studies that included images of the child’s maxillary central incisor teeth (19–21).

## 2. Methods

The study sample was mother and child dyads (n=161) who were a self-selected subset from the larger RCT and follow-up studies including dental imaging with detailed description previously published (19–21). From those data resources, we chose variables that had previously been associated with DDE as evidenced by publication (15, 22–30). We also included variables unique to our datasets that purposely included the period of 12 weeks of gestation through 4-6 weeks’ early infancy to encompass the developmental duration for the primary maxillary central incisor teeth. The variables included maternal predictors: mother’s age, pre-pregnancy body mass index (BMI), total count of months of antacid use from weeks 12-36; serum circulating levels of calcium (Ca), phosphorus (P); vitamin D as 25-hydroxyvitamin D (25(OH)D) and 1,25-dihydroxyvitamin D (1,25(OH)_2_D); and intact parathyroid hormone (iPTH). Child factors included gestational age, cord blood serum Ca, P, iPTH, 25(OH)D, and 1-25(OH)_2_D levels, and vitamin D binding protein (VDBP) genotype (focusing on 1s, 1f, and 2 genotypes as the three most common VDBP alleles). Early infancy diet was determined as whether or not the child had received formula by 4-6 weeks of age or was exclusively fed breastmilk. Child DDE outcomes were scored for the facial surfaces of the two maxillary central incisor teeth from digital images using the Enamel Defects Index (1). EH was operationalized by less enamel in the frontal (buccal-lingual) plane and PEB as less enamel in the median (mesial-distal) or the transverse (incisal-coronal) plane. Each facial surface was divided into 3 nonoverlapping regions (incisal, middle, and cervical) and the presence of each defect was scored for each region, resulting in child scores as status (binary) and extent (count, with a range of 0-6 per defect type for the 2 teeth per child) (21). Decayed, restored and missing teeth or regions (due to dental caries, trauma or exfoliation) were excluded from this study’s analyses. Intra-examiner agreement for EH was determined as “substantial” at the child level by a comparative rescore of the 6 tooth regions for 15% of the children (κ = 0.779) (31).

### 2.1 Variable Transformations

We log-transferred the mother’s BMI before model fitting. The functional vitamin D deficiency (FVDD) group was assigned for both the mothers during pregnancy and the children at birth. This criterion was applied based on previous studies that a concurrent maternal serum 25-dihydroxyvitamin D (25(OH)D) < 20ng/mL (deficiency) and intact parathyroid hormone (iPTH) > 65 pg/mL (abnormal; elevated) provided a definitive ratio i.e., ≤ 0.308 to identify FVDD (32). The mothers had monthly study visits from pregnancy week 12 through to delivery. Three time points during pregnancy were chosen for blood chemistry data because of their previously published trajectory of change: week 12 (starting point), week 28 (elbow point), and week 36 (ending point) (21). The child’s VDBP genotype was included as 6 levels (1s, 1f or 2 homozygous or 1s/1f, 1f/2 or 1s/2 heterozygous).

### 2.2 Bayesian Variable Selection

In our Bayesian paradigm, we used the Gibbs variable selection (GSV) with an indicator (π) to index the probability of including its corresponding predictor (33).

The linear function was:

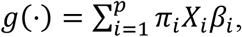

where *X*_*i*_ is the design matrix and *β*_*i*_ is the coefficient for the *i*^th^ predictor. The prior distribution of indicator was constructed in a hierarchical way:

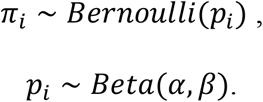

A uniform prior for the selection probability was chosen with *α* = *β* = 1.

For the binary outcomes, EH status, OP status, and PEB status, we applied the logistic regression with random effects as the following:

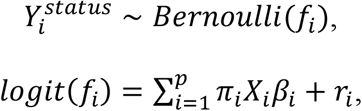

For the counted outcomes, EH extent, OP extent, and PEB extent, we applied the truncated Poisson regression with random effects as the following:

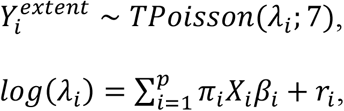

where 7 represents the upper limit of the count, and *B*_*i*_ is the random effect that allows for a random intercept for each subject under a normal prior distribution with mean 0 and variance *τ*^2^.

Following the variable selection, the final model was fitted again using only the selected predictors to obtain the corresponding coefficients.

### 2.3 Variable Imputation and Multinomial Regression

For missing values and outliers in some predictors, we assumed a prior distribution for each to impute values automatically when model fitting. The VDBP type was a multi-level categorical predictor (6 levels) and with some missing values. For the missing values we considered multinomial regression to impute its values with mother’s race as its predictor, which is based on previous studies about the association between race and genotype towards the VDBP types:

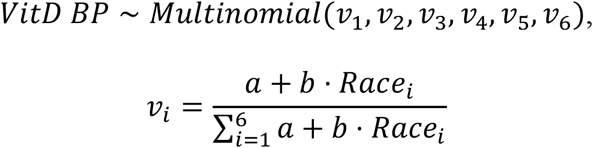

where *a* and *b* are the coefficients in the multinomial regression.

### 2.4 Fractional Polynomial Selection

After variable selection, we considered non-linear patterns by considering fractional polynomial models. The candidate power set was the commonly used:

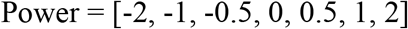

In particular, the power equals 0 means the logarithm transformation. We did the selection of power by comparing deviance information criterion (DIC), and the model with the smallest DIC value was chosen.

## 3. Results

### 3.1 Demographic information

Major characteristics of the 161 mothers who entered this study are presented in Table 1 by their treatment group assignment from the vitamin D supplementation RCT. The final mother-child dyad cohort sizes were 145 for EH, 144 for OP and 143 for PEB because of missingness in the children’s dental measurements, and also each dental defect had a different missing pattern.

**Table 1.**
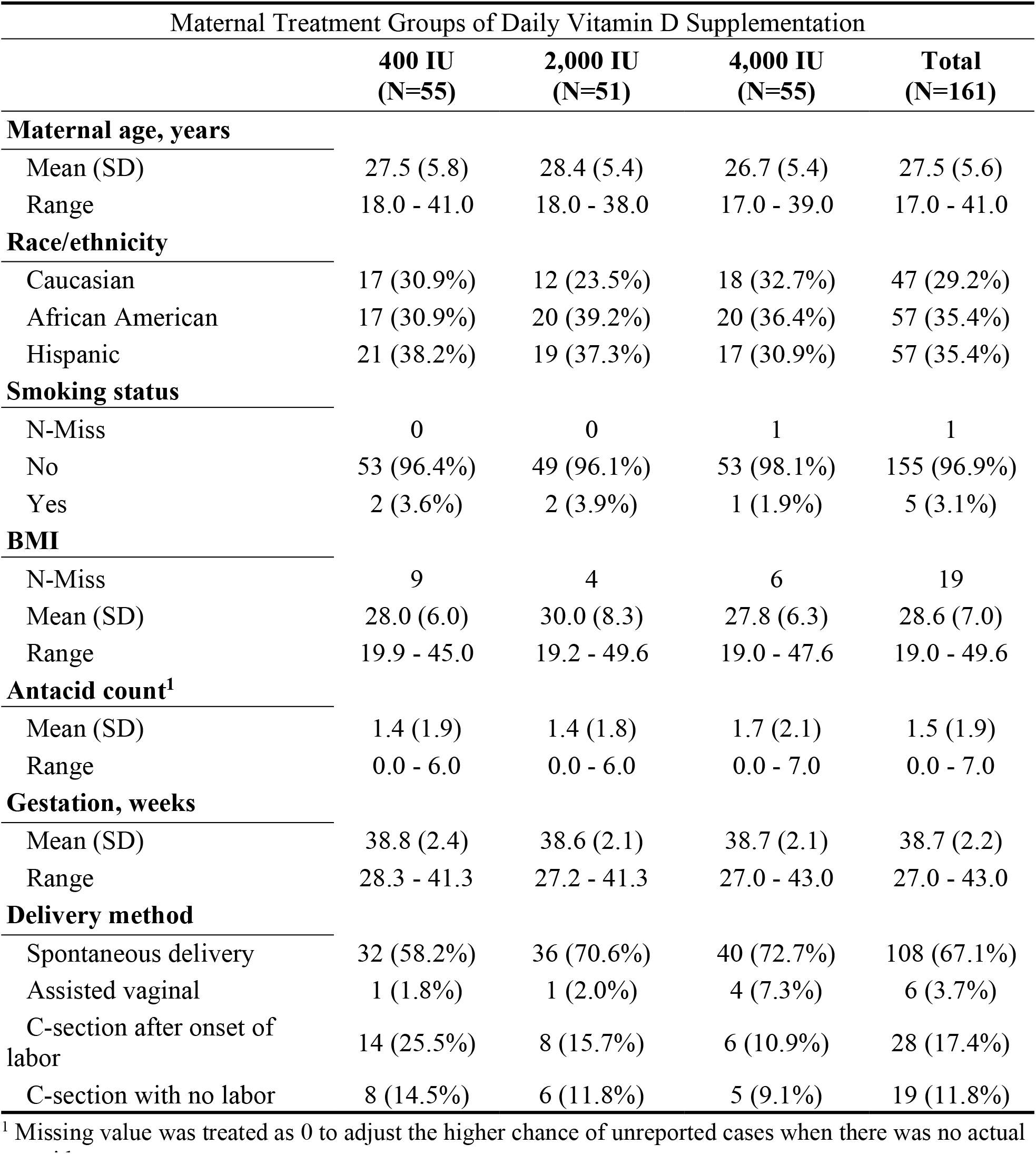
Maternal Socio-demographic, Behavioral and Delivery Characteristics by Treatment Group of the RCT of Daily Vitamin D Supplementation during Pregnancy

| Maternal Treatment Groups of Daily Vitamin D Supplementation |  |  |  |  |
| --- | --- | --- | --- | --- |
|  | 400 IU<br>(N=55) | 2,000 IU<br>(N=51) | 4,000 IU<br>(N=55) | Total<br>(N=161) |
| **Maternal age, years** |  |  |  |  |
| Mean (SD) | 27.5 (5.8) | 28.4 (5.4) | 26.7 (5.4) | 27.5 (5.6) |
| Range | 18.0 - 41.0 | 18.0 - 38.0 | 17.0 - 39.0 | 17.0 - 41.0 |
| **Race/ethnicity** |  |  |  |  |
| Caucasian | 17 (30.9%) | 12 (23.5%) | 18 (32.7%) | 47 (29.2%) |
| African American | 17 (30.9%) | 20 (39.2%) | 20 (36.4%) | 57 (35.4%) |
| Hispanic | 21 (38.2%) | 19 (37.3%) | 17 (30.9%) | 57 (35.4%) |
| **Smoking status** |  |  |  |  |
| N-Miss | 0 | 0 | 1 | 1 |
| No | 53 (96.4%) | 49 (96.1%) | 53 (98.1%) | 155 (96.9%) |
| Yes | 2 (3.6%) | 2 (3.9%) | 1 (1.9%) | 5 (3.1%) |
| **BMI** |  |  |  |  |
| N-Miss | 9 | 4 | 6 | 19 |
| Mean (SD) | 28.0 (6.0) | 30.0 (8.3) | 27.8 (6.3) | 28.6 (7.0) |
| Range | 19.9 - 45.0 | 19.2 - 49.6 | 19.0 - 47.6 | 19.0 - 49.6 |
| **Antacid count<sup>1</sup>** |  |  |  |  |
| Mean (SD) | 1.4 (1.9) | 1.4 (1.8) | 1.7 (2.1) | 1.5 (1.9) |
| Range | 0.0 - 6.0 | 0.0 - 6.0 | 0.0 - 7.0 | 0.0 - 7.0 |
| **Gestation, weeks** |  |  |  |  |
| Mean (SD) | 38.8 (2.4) | 38.6 (2.1) | 38.7 (2.1) | 38.7 (2.2) |
| Range | 28.3 - 41.3 | 27.2 - 41.3 | 27.0 - 43.0 | 27.0 - 43.0 |
| **Delivery method** |  |  |  |  |
| Spontaneous delivery | 32 (58.2%) | 36 (70.6%) | 40 (72.7%) | 108 (67.1%) |
| Assisted vaginal | 1 (1.8%) | 1 (2.0%) | 4 (7.3%) | 6 (3.7%) |
| C-section after onset of labor | 14 (25.5%) | 8 (15.7%) | 6 (10.9%) | 28 (17.4%) |
| C-section with no labor | 8 (14.5%) | 6 (11.8%) | 5 (9.1%) | 19 (11.8%) |

<sup>1</sup> Missing value was treated as 0 to adjust the higher chance of unreported cases when there was no actual antacid count.

**Table 2.** Children’s DDE by Status and by Extent

| DDE Observed Outcome Summary |  |  |  |  |
| --- | --- | --- | --- | --- |
|  | Status |  | Extent |  |
| **EH** | Counts | 60/145 | Mean (±SD) | 0.7 (±1.0) |
|  | Percentage | 41.4% | Range | 0 - 4 |
| **OP** | Counts | 45/144 | Mean (±SD) | 0.7 (±1.2) |
|  | Percentage | 31.2% | Range | 0 - 6 |
| **PEB** | Counts | 69/143 | Mean (±SD) | 0.8 (±1.0) |
|  | Percentage | 48.3% | Range | 0 - 5 |

### 3.2 Bayesian variable selection

To assess the importance of predictors in relation to the DDE outcomes, we employed Gibbs variable selection (33). The models with indicators greater than 0.5 were the optimal predictive models (34). We applied a slightly looser criterion (threshold at 0.3 in the first step and 0.4 in the following steps) to choose variables. We did this because the maternal blood chemistry predictors from 3 time points were used in our model and their mutual correlation could impact the indicator’s significance due to their mutual correlation. Variable selection results are shown in Table 3.

**Table 3.** The selected variables in the final models (Intercept is fixed for all models).

| Models | Variable Selection |
| --- | --- |
| EH Status | Intercept + mombmi<sup>1</sup> + momCa_28<sup>2</sup> |
| EH Extent | Intercept + mombmi<sup>1</sup> + momFVDD_28<sup>3</sup> |
| Opacity Status | Intercept + gestage<sup>4</sup> + childFVDD<sup>5</sup> + momFVDD_12<sup>6</sup> |
| Opacity Extent | Intercept + gestage<sup>4</sup> + childFVDD<sup>5</sup> |
| PEB Status | Intercept + mombmi<sup>1</sup> + momP_28<sup>7</sup> |
| PEB Extent | Intercept + mombmi<sup>1</sup> + momP_28<sup>7</sup> |

<sup>1</sup> mombmi: mother's BMI value at pre-pregnancy.
<sup>2</sup> momCa\_28: mother's calcium level at gestational week 28.
<sup>3</sup> momFVDD\_28: mother's FVDD condition at gestational week 28.
<sup>4</sup> gestage: gestational age in weeks at birth.
<sup>5</sup> childFVDD: child's FVDD condition at birth.
<sup>6</sup> momFVDD\_12: mother's FVDD condition at gestational week 12.
<sup>7</sup> momP\_28: mother's phosphorus level at gestational week 28.

The results for status models and extent score models for each defect are similar but not the same, except for the PEB outcome. We found the maternal FVDD was associated with both EH and OP outcomes, while the mother’s BMI was associated with EH and PEB outcomes, and the child’s gestational age week was only selected for the OP outcome.

As seen in Table 3, different time points (maternal weeks 12 and/or 28) for blood chemistries were selected for the models of the 3 outcomes. Moreover, for the OP models, the child’s FVDD and gestational age were selected as well as the maternal FVDD at week 12.

### 3.3 Effect of predictors

#### 3.3.1 Enamel Hyperplasia Defects

For EH status, the 95% credible interval for all the coefficients included 0, and the same results for EH extent model, but with a narrower interval width (see Appendix A2). The mean values of the coefficients were smaller than 0; namely, these predictors were negatively associated with the EH outcome, even though not dramatically under the 0.05 criteria.

#### 3.3.2 Opacity Defects

For OP status, the coefficient of gestational week had a 95% credible interval above 0, suggesting a positive association with the outcome, i.e., a longer gestation time was associated with a higher risk of having opacity defects in the child’s teeth. Holding others as constant, 1 unit increase in the gestation week increased the odds ratio as high as *B*^0.36^ = 1.43. However, the other predictors did not show an association with OP defects measured by binary status.

The OP extent score showed a strong association with gestational week based on the 95% credible interval. Holding others as constant, 1 unit increase in the gestation week increased the mean count (*λ*) as high as *B*^0.4^ = 1.49 time the baseline.

The child’s FVDD was negatively associated with OP extent score; however, the association only marginally approached the 0.05 level (Appendix A3), resulting in some overlaps between groups in Fig 2 (a). Note the ratio of 25(OH)D and iPTH below or equal to 0.308 indicated FVDD, which was then denoted as the FVDD group. We can see the FVDD group was associated with a higher risk of OP defects in child’s teeth. Holding others as constant, the FVDD group increased the average OP score (*λ*) as high as *B*^1.1^ = 3.00 times compared to the non-FVDD group.

**Figure 1.**
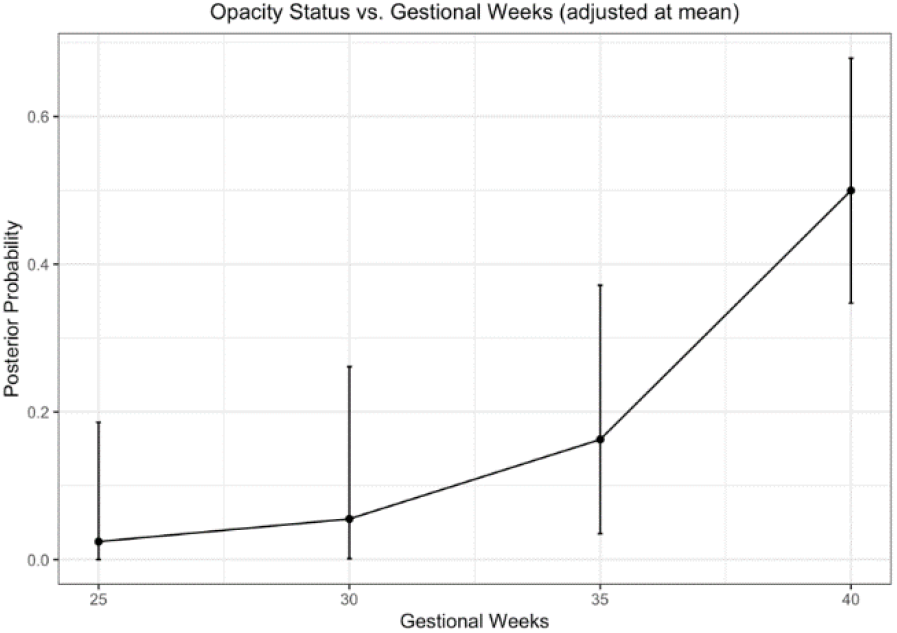
Posterior mean estimates of gestational weeks in OP status model with 95% credible interval.

**Figure 2.**
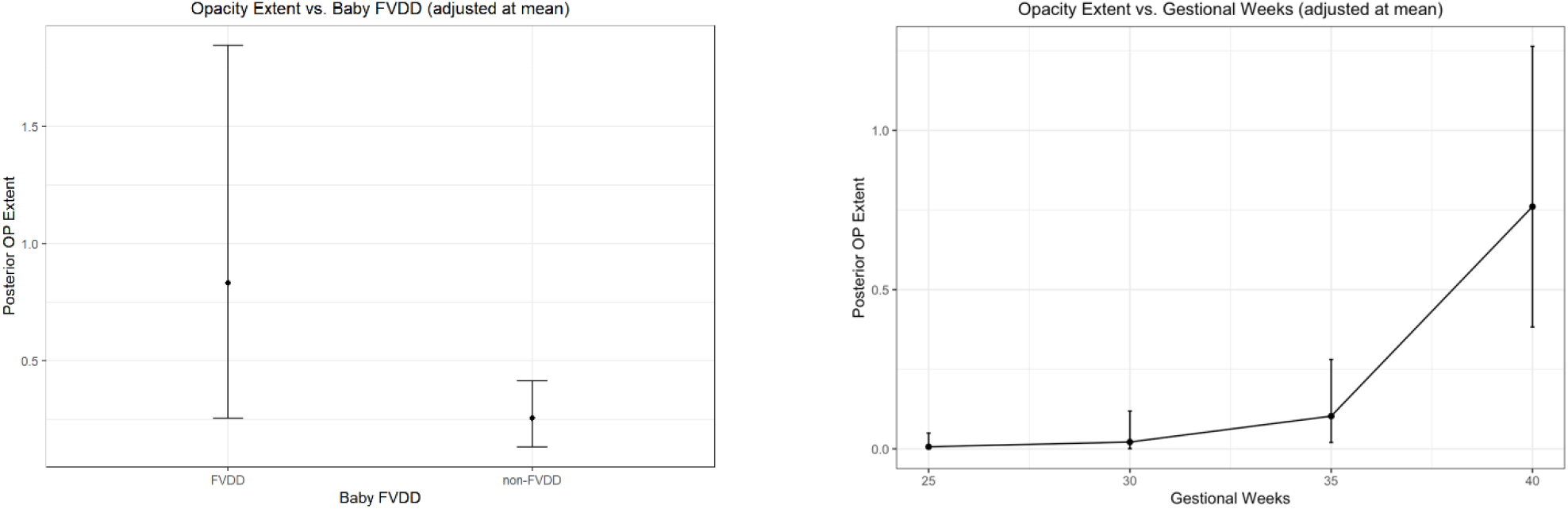
Posterior mean estimates (a: FVDD, b: gestational weeks) for OP extent model with 95% credible interval.

**Figure 3.**
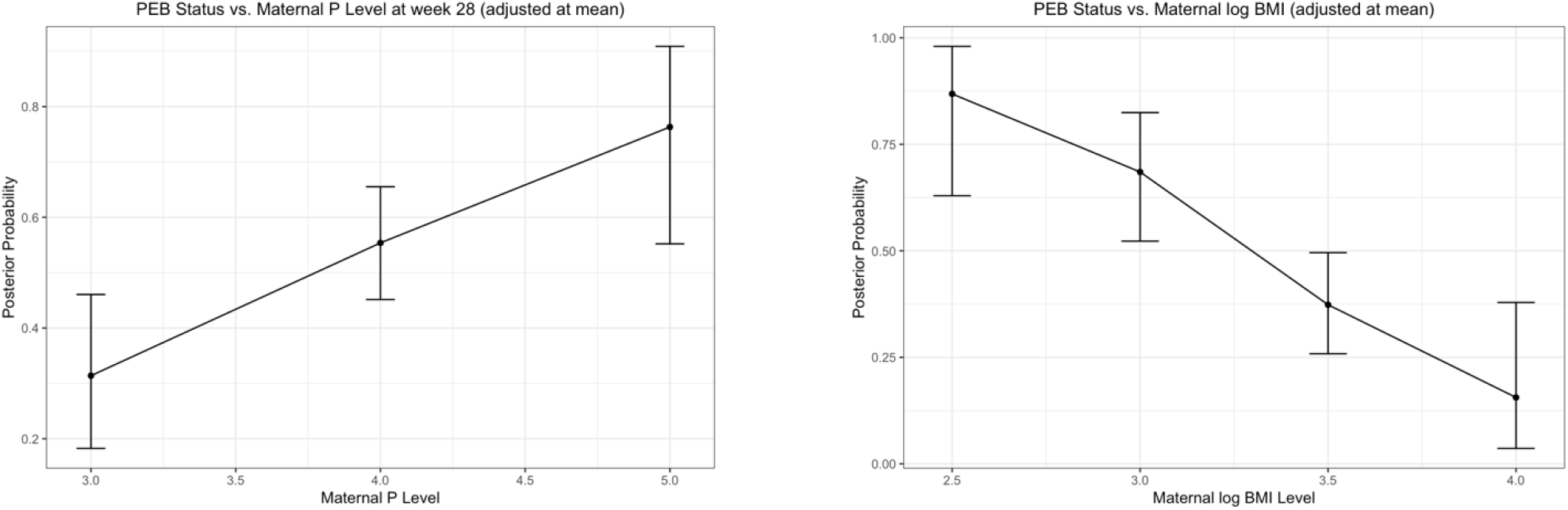
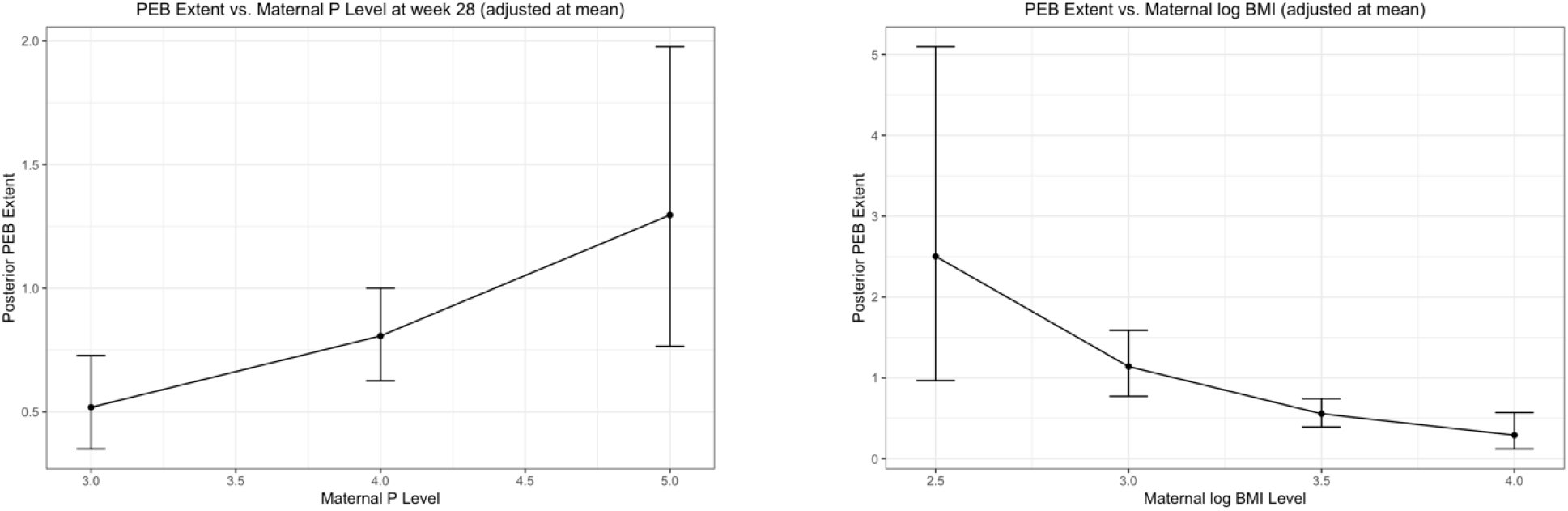
Posterior mean estimates (a: Phosphorus, b: BMI) for PEB status model with 95% credible interval, and posterior mean estimates (c: Phosphorus, d: BMI) for PEB extent model with 95% credible interval.

#### 3.3.3 Post Eruptive Breakdown Defects

For the PEB status, the coefficient of mother’s BMI was strongly and negatively associated with the outcome under a 95% credible interval. This indicates that a mother’s higher BMI value was associated with a lower risk of having PEB defects in child’s teeth. Holding others as constant, 1 unit increase from 21.5 in BMI decreased the odds ratio as much as *e*^−2.66×(log(22.5)−log(21.5))^ = 0.89.

The PEB score also showed a strong association with mother’s BMI. Holding others as constant, 1 unit increase from 21.5 in BMI decreased the average score (*λ*) as much as *e*^−1.43×(log(22.5)−log(21.5))^ = 0.93.

Moreover, the maternal serum P level was strongly and positively associated with both PEB status and extent score, according to the 95% credible interval. Holding others as constant, 1 unit increase in phosphorus increased the odds ratio as much as *e*^1.02^ = 2.77 for PEB status and increased the average score (*λ*) as much as 1.57. (Appendix A4).

Comparing status and extent models for each specific defect, the predictors showed the same direction towards the outcome on average, if they shared the same predictor set. Also, the coefficients for the extent models commonly had a shorter 95% credible interval width.

#### 3.4 Fractional Polynomial Models

For EH status models, the main effects selected were maternal BMI and maternal Ca at week 28. We tested the fractional polynomial (FP) powers on these two predictors. However, the minimum DIC was 202.8 (Appendix A5), which is trivially improved compared with the linear model (DIC = 205.4). Thus, we kept the original linear trend to consider interactions. For EH extent, the main effects selected were maternal BMI and maternal FVDD at week 28. The latter one was a categorical predictor, so we tested fractional polynomial on the maternal BMI only. Like the status model, DIC values indicated the linear trend is a good fit and we kept it due to simplicity. Similarly, the other four models - OP status, OP extent, PEB status and PEB extent - showed the non-linear terms were not necessary. Though non-linear trends were found by mean trajectory plots, e.g., OP status versus mother’s BMI, we kept the linear trend, for simplicity and explanation consideration.

#### 3.5 Models with Interaction terms

We checked the need to add interaction terms after the fractional polynomial selection. For all 6 parallel models we added possible interaction terms with the main effects fixed in the models. The coefficients of interaction terms all had a 95% credible interval including 0 (see Appendix A6). The DIC values for these models were slightly larger or the same compared with the models without interaction effects. Thus, we did not need to build the models with interaction terms.

## 4. Discussion

Our Bayesian model selection identified maternal and child predictors for the outcome of developmental defects of enamel (EH, OP and PEB) in the primary maxillary central incisor teeth. This unique study contributes knowledge to the etiology of these three major localized defects by having potential predictors that were periodically measured during the specific period of that tooth development. The key predictors for DDE were the mother’s pre-pregnancy BMI, her serum Ca and P levels at 28 weeks of pregnancy, and her FVDD condition at 12 and 28 weeks of pregnancy; and the child’s gestational age in weeks and FVDD at birth. The models for EH and PEB shared the predictor of mother’s pre-pregnancy BMI; whereas the OP model did not include those predictors. The mother’s FVDD at 12 weeks of pregnancy was important for OP and the mother’s FVDD at 28 weeks was important for EH.

New to this study was the predictor FVDD, determined as the ratio of 25(OH)D / iPTH ≤ 0.308 (20) at specific time points, namely at pregnancy weeks 12 and 28, and for the child at birth. This use of a FVDD grouping helped to reduce the number of predictors and also presented a clearer pattern that iPTH dominated the effect of 25(OH)D towards the defects, compared to our previous study that used 25(OH)D and iPTH levels only as independent variables (21). In future studies we will further assess FVDD predictability when used as a continuous variable over the weeks of pregnancy.

A strength of this observational cohort design using secondary analyses of existing data was the inclusion of maternal and child biomarkers of the key components of enamel formation e.g., Ca, P, 25(OH)D, and iPTH with biological plausibility for relation to enamel formation. Most all categories of previously identified high risk factors associated with these dental defects were included. We assessed early infancy feeding habits as exclusively breast milk or formula fed by 4-6 weeks. Our sample included African American, Caucasian, and Hispanic mothers; and the child’s genotype for VDBP. The self-selected sample for participation through to the dental imaging was from a population of healthy mothers that excluded any conditions related to vitamin D regulation (19). And yet in this healthy sample we found DDE rates of 41.4% for EH, 31.2% for OP and 48.3% for PEB. Our defect rates are not comparable to most published data because our mothers and children were very healthy and the outcome measures were restricted to only 2 teeth (PMCI), rather than the full primary dentition. Also, there is the potential confounding in the mothers because of participation in an RCT of vitamin D supplementation. However, the mothers’ characteristics by treatment group were similar and also our key variables were from circulating blood chemistries rather than vitamin D intake.

In consideration of sample size and the impact of outliers, we found that the risk of OP changed slowly when the gestational age was less than 36 weeks, and then the risk for OP increased dramatically beyond 36 weeks. Thus, we are more likely to state the effect of gestational age as positive when the value of gestational week is relatively very high. Similar ideas related to the effect of outliers may also explain the impact of the mother’s BMI and serum chemistry predictors. One explanation is that the range of our predictors were concentrated within a certain interval, making their extreme values to have a greater leverage effect on the results. This may be due to a relatively limited sample size, as well as limited heterogeneity in the healthy cohort.

Our methodology and results provide a roadmap for assessing time appropriate exposure measures during specific tooth development to better understand the etiology of developmental defects of enamel. Though the PMCI are present at birth with the structural abnormalities of EH and OP, these defects are not recognized until after the teeth erupt into the oral cavity. Our ongoing studies continue to examine the interrelationships among the three defects and their exposome to better understand the *in utero* and early infancy development of these dental defects in the primary maxillary central incisor teeth. Our efforts continue towards improving the understanding of the development of enamel defects and possibly modifiable variables for the future prevention of DDE.

## Data Availability

All data produced in the present work are available at the completion of the ongoing research and upon reasonable request to the authors.

## Appendix

### A1. Results of Laboratory Measurements

**Table 1.** EH status versus children’s cord blood chemistry measurements.

| EH Status | 0 (N=85) | 1 (N=60) | Total (N=145) |
| --- | --- | --- | --- |
| **Total 25(OH)D (ng/mL)** |  |  |  |
| N-Missing | 3 | 4 | 7 |
| Mean (SD) | 22.58 (10.53) | 22.66 (10.37) | 22.62 (10.43) |
| Range | 3.60 - 52.00 | 5.30 - 45.50 | 3.60 - 52.00 |
| **1,25(OH)<sub>2</sub>D (pg/mL)** |  |  |  |
| N-Missing | 33 | 25 | 58 |
| Mean (SD) | 34.51 (12.82) | 40.63 (15.94) | 36.97 (14.39) |
| Range | 9.20 - 67.10 | 16.00 - 100.10 | 9.20 - 100.10 |
| **iPTH (pg/mL)** |  |  |  |
| N-Miss | 10 | 16 | 26 |
| Mean (SD) | 10.47 (9.85) | 6.66 (5.61) | 9.06 (8.70) |
| Range | 0.90 - 46.70 | 0.90 - 25.60 | 0.90 - 46.70 |
| **Ca (mg/dL)** |  |  |  |
| N-Missing | 11 | 14 | 25 |
| Mean (SD) | 10.05 (0.57) | 10.07 (0.68) | 10.06 (0.61) |
| Range | 8.00 - 11.40 | 6.90 - 11.50 | 6.90 - 11.50 |
| **P (mg/dL)** |  |  |  |
| N-Missing | 12 | 18 | 30 |
| Mean (SD) | 6.11 (1.09) | 5.81 (1.26) | 6.00 (1.16) |
| Range | 4.20 - 10.20 | 0.50 - 8.50 | 0.50 - 10.20 |

| **EH Status** |  |  |  |
| --- | --- | --- | --- |
|  | 0 (N=85) | 1 (N=60) | Total (N=145) |
| **VDBP Genotype** |  |  |  |
| N-Missing | 35 | 17 | 52 |
| 1F/1F | 13 (26.0%) | 10 (23.3%) | 23 (24.7%) |
| 1F/2 | 2 (4.0%) | 7 (16.3%) | 9 (9.7%) |
| 1S/1F | 13 (26.0%) | 8 (18.6%) | 21 (22.6%) |
| 1S/1S | 8 (16.0%) | 9 (20.9%) | 17 (18.3%) |
| 1S/2 | 11 (22.0%) | 7 (16.3%) | 18 (19.4%) |
| 2/2 | 3 (6.0%) | 2 (4.7%) | 5 (5.4%) |

Note: Children's laboratory and chemical results across EH status, which have a smaller cohort size (n=145) due to missingness of study follow-up, and the unidentifiable defect conditions during diagnosis.

**Table 2.** Opacity status versus children’s cord blood chemistry measurements.

| **Opacity Status** |  |  |  |
| --- | --- | --- | --- |
|  | 0 (N=99) | 1 (N=45) | Total (N=144) |
| **25(OH)D (ng/mL)** |  |  |  |
| N-Missing | 5 | 2 | 7 |
| Mean (SD) | 22.57 (10.52) | 22.73 (10.47) | 22.62 (10.47) |
| Range | 3.60 - 52.00 | 3.60 - 47.80 | 3.60 - 52.00 |
| **1,25(OH)<sub>2</sub>D (pg/mL)** |  |  |  |
| N-Missing | 39 | 19 | 58 |
| Mean (SD) | 38.88 (15.62) | 32.14 (9.93) | 36.84 (14.42) |
| Range | 9.20 - 100.10 | 16.00 - 55.30 | 9.20 - 100.10 |
| **iPTH (pg/mL)** |  |  |  |
| N-Missing | 16 | 9 | 25 |

**Opacity Status**
|  | 0 (N=99) | 1 (N=45) | Total (N=144) |
| --- | --- | --- | --- |
| Mean (SD) | 9.43 (9.08) | 8.40 (7.90) | 9.12 (8.72) |
| Range | 0.90 - 46.70 | 0.90 - 41.90 | 0.90 - 46.70 |
| **Ca (mg/dL)** |  |  |  |
| N-Missing | 11 | 14 | 25 |
| Mean (SD) | 10.07 (0.65) | 10.03 (0.51) | 10.06 (0.61) |
| Range | 6.90 - 11.50 | 8.50 - 11.20 | 6.90 - 11.50 |
| **P (mg/dL)** |  |  |  |
| N-Missing | 14 | 15 | 29 |
| Mean (SD) | 6.08 (1.08) | 5.73 (1.33) | 5.99 (1.16) |
| Range | 3.90 - 10.20 | 0.50 - 7.50 | 0.50 - 10.20 |
| **VDBP Genotype** |  |  |  |
| N-Missing | 35 | 17 | 52 |
| 1F/1F | 16 (25.0%) | 7 (25.0%) | 23 (25.0%) |
| 1F/2 | 7 (10.9%) | 2 (7.1%) | 9 (9.8%) |
| 1S/1F | 16 (25.0%) | 5 (17.9%) | 21 (22.8%) |
| 1S/1S | 11 (17.2%) | 5 (17.9%) | 16 (17.4%) |
| 1S/2 | 11 (17.2%) | 7 (25.0%) | 18 (19.6%) |
| 2/2 | 3 (4.7%) | 2 (7.1%) | 5 (5.4%) |

Note: Children's laboratory and chemical results across OP status, which have a smaller cohort size (n=144) due to missingness of study follow-up, and the unidentifiable defect conditions during diagnosis.

**Table 3.**
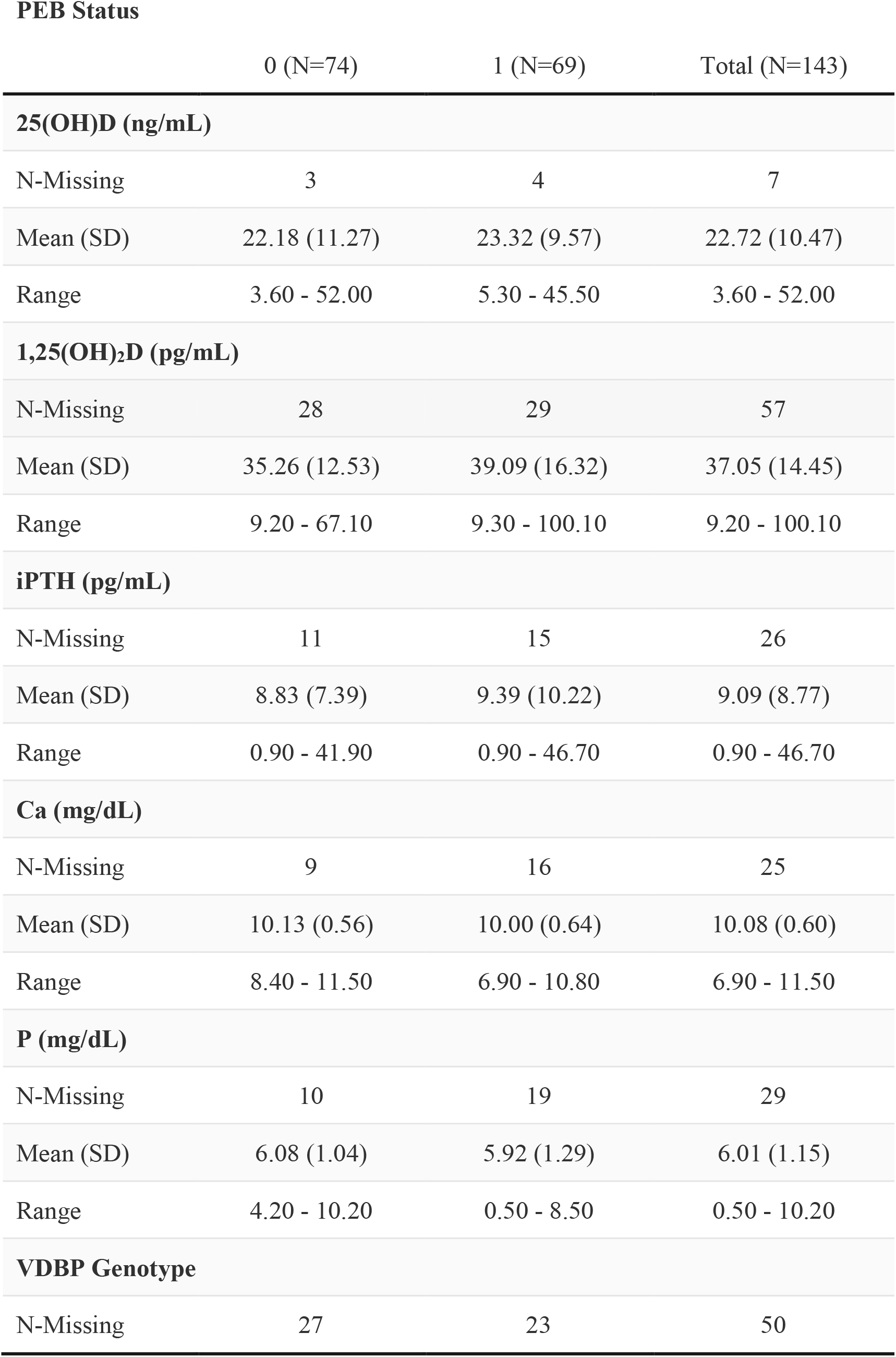
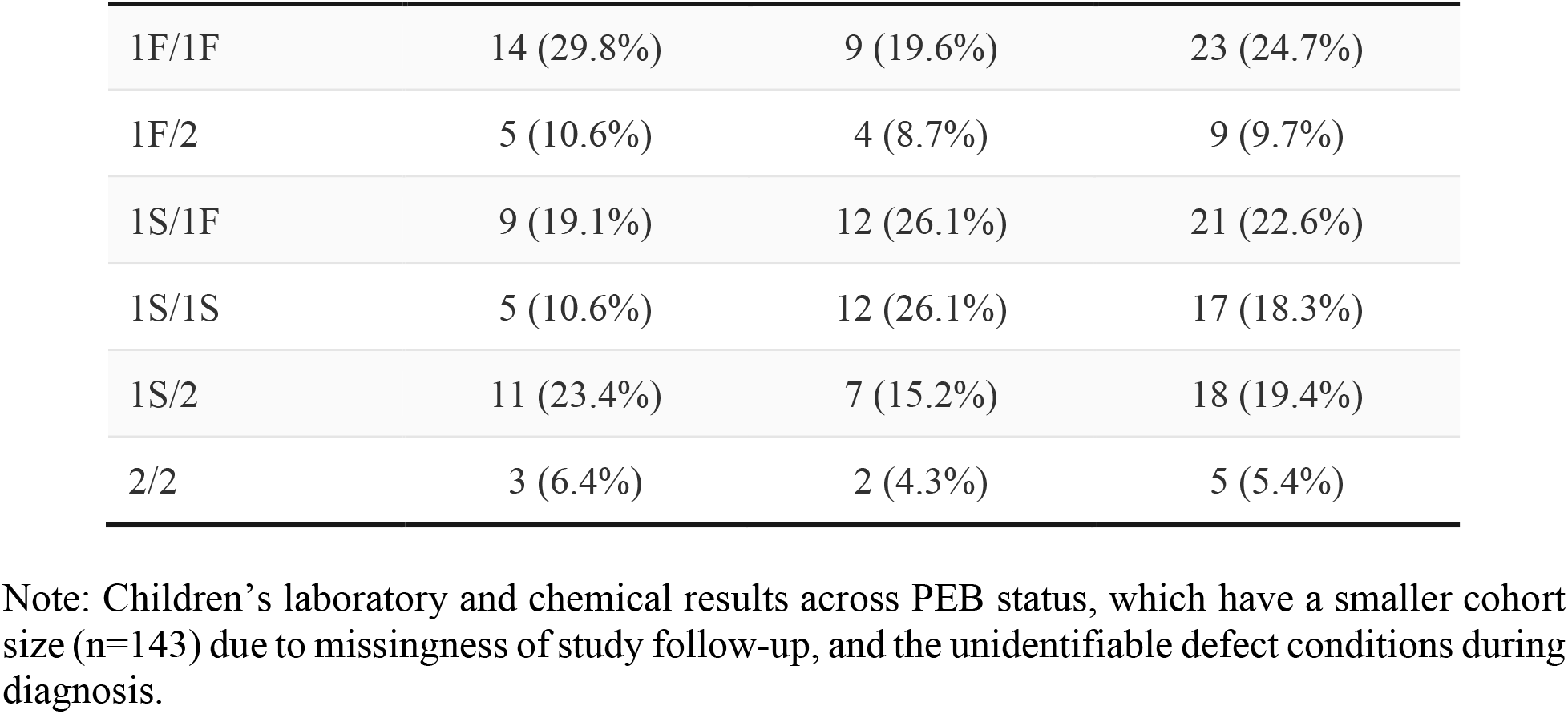
PEB status versus children’s cord blood chemistry measurements.

### A2. Model Coefficients for EH

A2. Model Coefficients for EH EH Status: Statistics of Coefficients
| Predictors | Mean | SD | 2.5% | 50% | 97.5% |
| --- | --- | --- | --- | --- | --- |
| **mombmi** | -0.95 | 0.86 | -2.76 | -0.89 | 0.46 |
| **momCa_28** | -0.53 | 0.52 | -1.64 | -0.48 | 0.36 |
| **Intercept** | -0.33 | 0.18 | -0.69 | -0.33 | 0.01 |
| **Deviance** | 281.20 | 15.56 | 239.30 | 284.60 | 300.80 |

EH Extent Score: Statistics of Coefficients
| Predictors | Mean | SD | 2.5% | 50% | 97.5% |
| --- | --- | --- | --- | --- | --- |
| **mombmi** | -0.8 | 0.6 | -2.0 | -0.8 | 0.2 |
| **momFVDD_28** | -0.5 | 0.4 | -1.4 | -0.5 | 0.2 |
| **Intercept** | -0.1 | 0.4 | -0.9 | -0.1 | 0.8 |
| **Deviance** | 3146.0 | 17.8 | 3112.0 | 3146.0 | 3180.0 |

Note: The coefficients of the predictors for EH models under the final Bayesian model. There is no strong evidence for any predictor to be apart from 0 (for both status and extent score models), according to the 95% credible interval.

### A3. Model Coefficients for Opacity

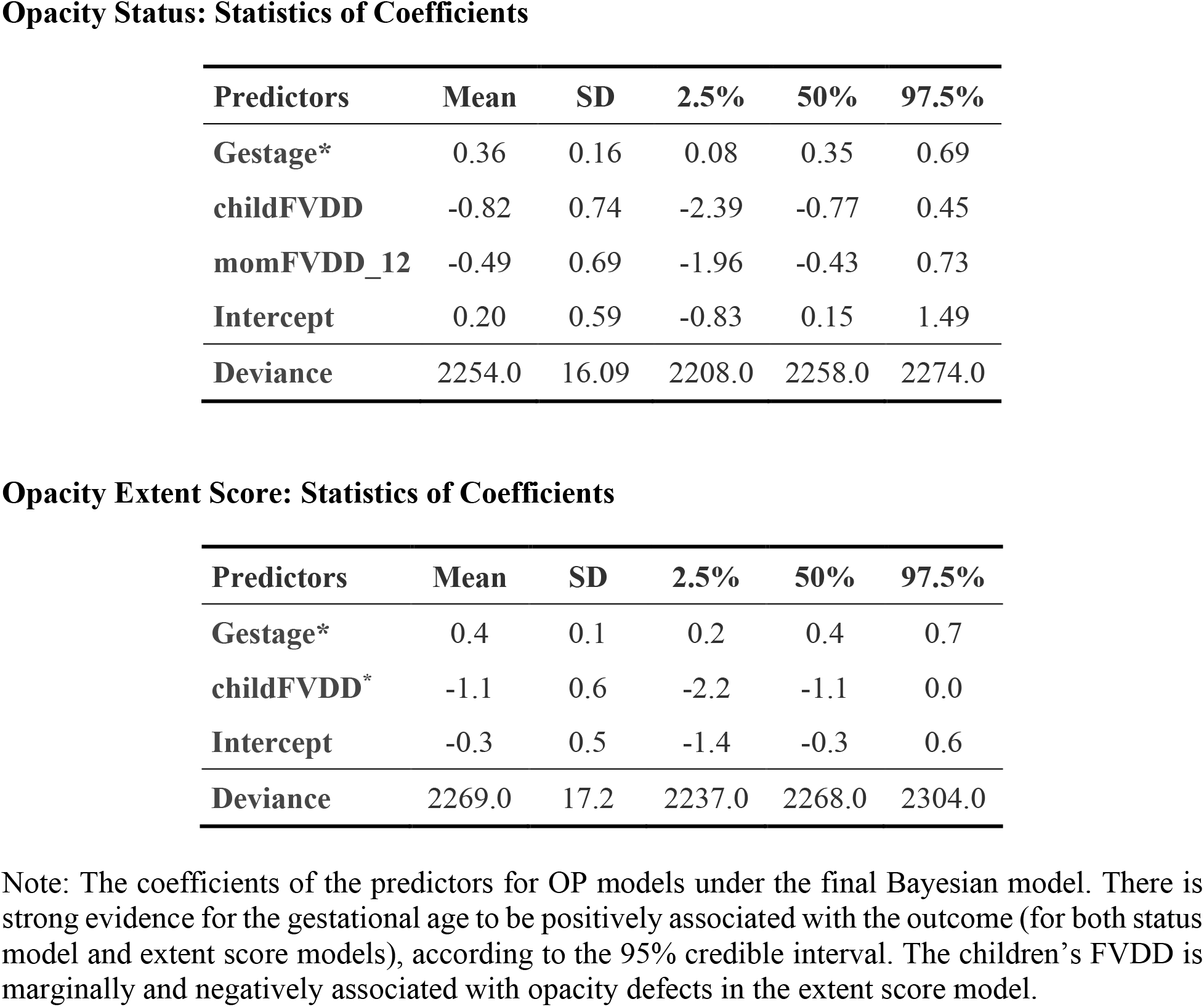

### A4. Model Coefficients for PEB

A4. Model Coefficients for PEB PEB Status: Statistics of Coefficients
| Predictors | Mean | SD | 2.5% | 50% | 97.5% |
| --- | --- | --- | --- | --- | --- |
| **mombmi\*** | -2.66 | 1.02 | -4.75 | -2.62 | -0.74 |
| **momP_28\*** | 1.02 | 0.39 | 0.27 | 1.01 | 1.83 |
| **Intercept** | -0.06 | 0.18 | -0.43 | -0.06 | 0.31 |
| **Deviance** | 398.60 | 14.91 | 359.70 | 401.40 | 418.80 |

PEB Extent Score: Statistics of Coefficients
| Predictors | Mean | SD | 2.5% | 50% | 97.5% |
| --- | --- | --- | --- | --- | --- |
| **mombmi\*** | -1.43 | 0.53 | -2.45 | -1.43 | -0.38 |
| **momP_28\*** | 0.45 | 0.18 | 0.09 | 0.46 | 0.79 |
| **Intercept\*** | -0.35 | 0.12 | -0.59 | -0.35 | -0.14 |
| **Deviance** | 536.40 | 11.49 | 511.50 | 537.10 | 557.00 |

Note: The coefficients of the predictors for PEB models under the final Bayesian model. There is strong evidence for mother's BMI and mother's phosphorous concentration (at week 28) to be different from 0 (for both status model and extent score models), according to the 95% credible interval. The intercept for the extent score model is even more strongly negative, which is the baseline under centered predictors.

### A5. DIC Values for Fractional Polynomial Selection

| Enamel Hyperplasia |  |  |  |  |  |  |  | Extent<sup>2</sup> |
| --- | --- | --- | --- | --- | --- | --- | --- | --- |
| Status<sup>1</sup> |  |  |  |  |  |  |  |  |
|  | -2 | -1 | -0.5 | log | 0.5 | 1 | 2 |  |
| -2 | 204.0 | 203.9 | 204.6 | 204.1 | 203.9 | 204.0 | 203.8 | 183.4 |
| -1 | 204.1 | 206.6 | 204.9 | 204.3 | 203.6 | 204.2 | 203.8 | 182.1 |
| -0.5 | 204.4 | 204.3 | 205.3 | 204.0 | 207.6 | 203.6 | 202.9 | 182.4 |
| log | 203.7 | 203.6 | 206.6 | 205.6 | 204.8 | 205.1 | 204.7 | 190.5 |
| 0.5 | 203.0 | 204.8 | 205.4 | 203.9 | 204.2 | 206.1 | 204.5 | 182.1 |
| 1 | 203.6 | 203.7 | 204.1 | 204.1 | 206.1 | 205.4 | 203.9 | 179.5 |
| 2 | 202.8 | 204.8 | 205.5 | 205.8 | 204.4 | 204.8 | 205.1 | 176.7 |

| Opacity Defects |  |  |
| --- | --- | --- |
| Status<sup>3</sup> |  | Extent<sup>3</sup> |
| -2 | 183.4 | 277.3 |
| -1 | 182.1 | 275.9 |
| -0.5 | 182.4 | 275.5 |
| log | 190.5 | 276.1 |
| 0.5 | 182.1 | 276.5 |
| 1 | 179.5 | 276.2 |
| 2 | 176.7 | 274.9 |

| Post Eruptive Break Defects |  |  |  |  |  |  |  |  |  |  |  |  |  |  |
| --- | --- | --- | --- | --- | --- | --- | --- | --- | --- | --- | --- | --- | --- | --- |
| Status<sup>4</sup> |  |  |  |  |  |  |  | Extent<sup>4</sup> |  |  |  |  |  |  |
|  | -2 | -1 | -0.5 | log | 0.5 | 1 | 2 | -2 | -1 | -0.5 | log | 0.5 | 1 | 2 |
| -2 | 205.1 | 203.9 | 207.3 | 204.5 | 205.5 | 204.0 | 200.6 | 353.3 | 351.9 | 352.9 | 354.9 | 355.5 | 354.3 | 353.0 |
| -1 | 203.2 | 203.3 | 204.1 | 204.0 | 205.1 | 202.9 | 201.0 | 353.3 | 350.9 | 350.0 | 355.0 | 355.7 | 354.3 | 353.1 |
| -0.5 | 203.2 | 201.1 | 202.6 | 203.8 | 205.2 | 202.9 | 200.6 | 353.8 | 351.9 | 351.2 | 355.1 | 356.0 | 354.4 | 353.4 |
| log | 208.7 | 206.0 | 204.4 | 191.2 | 189.0 | 191.5 | 195.1 | 353.3 | 353.5 | 354.3 | 346.5 | 345.2 | 347.3 | 348.9 |
| 0.5 | 203.2 | 203.9 | 203.6 | 189.9 | 190.0 | 189.7 | 194.5 | 353.9 | 353.7 | 354.7 | 348.6 | 347.3 | 349.1 | 350.1 |
| 1 | 202.5 | 200.5 | 201.1 | 191.9 | 193.3 | 190.6 | 189.8 | 352.8 | 353.1 | 353.8 | 345.5 | 344.0 | 345.9 | 347.6 |
| 2 | 199.7 | 200.1 | 200.5 | 195.1 | 197.2 | 194.4 | 192.5 | 350.5 | 350.6 | 351.5 | 343.9 | 344.3 | 343.8 | 344.7 |

<sup>1</sup> Fractional polynomial selection between maternal BMI (in rows) and Ca level at week 28 (in columns).
<sup>2</sup> Fractional polynomial selection of maternal BMI (in rows).
<sup>3</sup> Fractional polynomial selection of gestational week (in rows).
<sup>4</sup> Fractional polynomial selection between maternal BMI (in rows) and P level at week 28 (in columns).

### A6. Coefficients of Models with Interaction Terms

| EH Status: Statistics of Coefficients |  |  |  |  |  |
| --- | --- | --- | --- | --- | --- |
|  | Mean | SD | 2.5% | 50% | 97.5% |
| **mombmi\*momCa_28** | -0.08 | 0.64 | -1.44 | -0.06 | 1.15 |
| **mombmi** | -0.93 | 0.85 | -2.76 | -0.86 | 0.48 |
| **momCa_28** | -0.53 | 0.52 | -1.65 | -0.48 | 0.36 |
| **Intercept** | -0.33 | 0.18 | -0.69 | -0.32 | 0.02 |

| EH Extent: Statistics of Coefficients |  |  |  |  |  |
| --- | --- | --- | --- | --- | --- |
|  | Mean | SD | 2.5% | 50% | 97.5% |
| **mombmi\*momFVDD_28** | -0.05 | 0.61 | -1.22 | -0.06 | 1.19 |
| **mombmi** | -0.82 | 0.81 | -2.48 | -0.76 | 0.58 |
| **momFVDD_28** | -0.54 | 0.42 | -1.38 | -0.53 | 0.23 |
| **Intercept** | -0.09 | 0.40 | -0.85 | -0.09 | 0.70 |

| Opacity Status: Statistics of Coefficients |  |  |  |  |  |
| --- | --- | --- | --- | --- | --- |
|  | Mean | SD | 2.5% | 50% | 97.5% |
| **gestage\*childFVDD** | 0.0000 | 0.0022 | -0.0038 | -0.0002 | 0.0045 |
| **gestage\*momFVDD_12** | 0.0016 | 0.0022 | -0.0022 | 0.0015 | 0.0062 |
| **gestage** | 0.0020 | 0.0015 | -0.0005 | 0.0019 | 0.0058 |
| **childFVDD** | -2.7220 | 2.8950 | -9.3230 | -2.4310 | 1.8340 |
| **momFVDD_12** | -3.6780 | 3.0860 | -9.9580 | -3.4130 | 1.3210 |
| **Intercept** | -0.1810 | 0.8555 | -1.9000 | -0.0981 | 1.1350 |

| Opacity Extent: Statistics of Coefficients |  |  |  |  |  |
| --- | --- | --- | --- | --- | --- |
|  | Mean | SD | 2.5% | 50% | 97.5% |
| **gestage\*childFVDD** | 0.1691 | 0.4527 | -0.7531 | 0.1795 | 1.1080 |
| **gestage** | 0.2724 | 0.4595 | -0.6475 | 0.2388 | 1.2260 |
| **childFVDD** | -1.1150 | 0.5825 | -2.2630 | -1.1220 | 0.0203 |
| **Intercept** | -0.2915 | 0.4939 | -1.3230 | -0.2578 | 0.6148 |

| PEB Status: Statistics of Coefficients |  |  |  |  |  |
| --- | --- | --- | --- | --- | --- |
|  | Mean | SD | 2.5% | 50% | 97.5% |
| **mombmi\*momP_28** | -0.08 | 0.59 | -1.32 | -0.07 | 1.06 |
| **mombmi** | -2.62 | 1.02 | -4.76 | -2.59 | -0.68 |
| **momP_28** | 1.01 | 0.38 | 0.29 | 1.01 | 1.77 |
| **Intercept** | -0.06 | 0.18 | -0.42 | -0.06 | 0.29 |

| PEB Extent: Statistics of Coefficients |  |  |  |  |  |
| --- | --- | --- | --- | --- | --- |
|  | Mean | SD | 2.5% | 50% | 97.5% |
| **mombmi\*momP_28** | -0.11 | 0.49 | -1.09 | -0.10 | 0.86 |
| **mombmi** | -1.41 | 0.53 | -2.45 | -1.40 | -0.35 |
| **momP_28** | 0.45 | 0.18 | 0.10 | 0.45 | 0.80 |
| **Intercept** | -0.36 | 0.12 | -0.60 | -0.35 | -0.14 |

For all the interaction term added in the models, the 95% credible intervals included 0 for the interaction coefficients.

